# Validation of the RT-LAMP assay in a large cohort of nasopharyngeal swab samples shows that it is a useful screening method for detecting SARS-CoV-2 and its VOC variants

**DOI:** 10.1101/2022.02.15.22270954

**Authors:** Mireya Cisneros-Villanueva, Sugela Blancas, Alberto Cedro-Tanda, Magdalena Ríos-Romero, Eduardo Hurtado-Córdova, Oscar Almaraz-Rojas, Diana R. Ortiz-Soriano, Víctor Álvarez-Hernández, Ivonne E. Arriaga-Guzmán, Laura Tolentino-García, Antonia Sánchez-Vizcarra, Laura F. Lozada-Rodríguez, Irlanda Peralta-Arrieta, José E. Pérez-Aquino, Marco A. Andonegui-Elguera, Mariana Cendejas-Orozco, Alfredo Mendoza-Vargas, Juan P. Reyes-Grajeda, Abraham Campos-Romero, Jonathan Alcantar-Fernández, José Luis Moreno-Camacho, Jorge Gallegos-Rodriguez, Marco Esparza-Luna-Ruiz, Jesus Ortiz-Ramirez, Mariana Benitez Gonzalez, Laura Uribe-Figueroa, Rosaura Ruiz, Ofelia Angulo, Luis A. Herrera, Alfredo Hidalgo-Miranda

**Affiliations:** Laboratorio de Genómica del Cáncer, Instituto Nacional de Medicina Genómica, INMEGEN, Mexico City, Mexico; Cátedras CONACYT - Laboratorio de Genómica del Cáncer, Instituto Nacional de Medicina Genómica, INMEGEN, Mexico City, Mexico; Instituto Nacional de Medicina Genómica, INMEGEN, Mexico City, Mexico; Innovation and Research Department, Salud Digna, Sinaloa, Mexico; Clinical Laboratory Division, Salud Digna, Sinaloa, Mexico; Hospital General Ajusco Medio, Mexico City, Mexico; Arion Genética, Mexico City, Mexico; Secretaría de Educación, Ciencia, Tecnología e Innovación de la Ciudad deMexico; Unidad de Investigación Biomédica en Cáncer, Instituto Nacional de Cancerología-Instituto de Investigaciones Biomédicas, UNAM

**Keywords:** COVID-19, SARS-CoV-2, RT-LAMP assay, validation, RT-qPCR, nasopharyngeal swab, VOC, VOI, VUM, FMV, variants

## Abstract

The COVID-19 pandemic is challenging the global supply chain and equipment needed for mass testing with RT-qPCR, the gold standard for SARS-CoV-2 diagnosis. Here, we propose the RT-LAMP assay as an additional strategy for rapid virus diagnosis. However, its validation as a diagnostic method remains uncertain. In this work, we validated the RT-LAMP assay in 1,266 nasopharyngeal swab samples with confirmed diagnosis by CDC 2019-nCoV RT-qPCR. Our cohort was divided, the first (n=984) was used to evaluate two sets of oligonucleotides (S1 and S3) and the second (n=281) to determine whether RT-LAMP could detect samples with several types of variants. This assay can identify positive samples by color change or fluorescence within 40 minutes and shows high concordance with RT-qPCR in samples with CT ≤35. Also, S1 and S3 are able to detect SARS-CoV-2 with a sensitivity of 68.4% and 65.8%, and a specificity of 98.9% and 97.1%, respectively. Furthermore, RT-LAMP assay identified 279 sequenced samples as positive (99.3% sensitivity) corresponding to the Alpha, Beta, Gamma, Delta, Epsilon, Iota, Kappa, Lambda, Mu and Omicron variants. In conclusion, RT-LAMP is able to identify SARS-CoV-2 with good sensitivity and excellent specificity, including all VOC, VOI, VUM and FMV variants.

## Introduction

The current COVID-19 pandemic caused by infection with the SARS-CoV-2 coronavirus continues to be a global challenge. On January 30, 2020, the World Health Organization (WHO) declared the COVID-19 outbreak an international public health emergency in accordance with the International Health Regulations (1). According to WHO, the number of new COVID-19 cases increased over the past week (31 January to 6 February 2022), with more than 392 million confirmed cases and over 5.7 million deaths have been reported worldwide (new deaths increased by 7%) (2), demonstrating how swiftly the virus spreads and the need to continue with rapid detection of positive cases as well as vaccination programs. Many countries have focused on their national vaccination programs, even though the COVID-19 vaccines have shown to be safe and effective against COVID-19, manufacturing and distribution issues persist (3).

On the other hand, it is well known that the biological variability of the SARS-CoV-2 coronavirus represents a challenge, as the virus naturally mutates to form new variants during transmission, making it difficult to determine their impact on the current disease pathology, such as higher infectivity, which could affect immune response and vaccine design (4-7). Several SARS-CoV-2 strains have been sequenced to date, and diverse viral genomic changes have begun to emerge, resulting in new variant strains, such as the Delta variant (B.1.617.2), which is highly contagious, evades immunity better than existing variants, causes severe disease, and is more resistant to preventive measures, treatments, and vaccines (7-10).

Recently, the Omicron variant has been described to be phylogenetically distinct from other variants and it is highly transmissible and rapidly spreading, with an average doubling time of two days. Although prior infection and immunization offer little or no protection against infection with omicron, they do appear to protect against hospitalization and severe disease, in a similar fashion to the delta variant. Booster vaccines have had no discernible impact on omicron’s spread (11, 12). For a variety of reasons, the total risk associated with Omicron remains remarkably high. According to current data, Omicron has a significant growth advantage over Delta, resulting in rapid community spread. This rapid increase in cases will result in more hospitalizations, putting excessive stress on health-care systems and causing severe morbidity, especially among vulnerable patients (13). Hence, the protection provided by vaccines against future variants is uncertain, and outbreaks may reappear (14). Thus, recommendations for national testing strategies and the use of PCR and rapid tests in different transmission scenarios of the COVID-19 outbreak are required for early identification of infected individuals and pandemic control (15) particularly in countries with few or no SARS-CoV-2 diagnostic tests and low vaccination rates.

In this context, COVID-19 pandemic has challenged the worldwide chain of supplies for the reagents and equipment needed for mass testing. The deployment of diagnostic tests is required to manage the sanitary emergency, which has proven problematic, particularly in locations where access to testing facilities capable of processing copious amounts of samples is difficult. This scenario, combined with a global scarcity of testing reagents and equipment, is the major constraint for implementing massive sampling and testing, which in turn, is required for proper patient management, and an effective mitigation and surveillance actions (16).

The CDC designed the 2019-nCoV CDC kit, which uses N1 and N2 probes to identify SARS-CoV-2 and RNase P as an RNA extraction quality control as well as additional protocols based on RT-qPCR, which is the gold standard for virus detection worldwide (17-22). However, implementation of the “gold standard” has several limitations. First, the test takes several hours to complete and requires extensive human labor. Second, resources such as RNA extraction kits had become scarce, and third, qPCR machines availability had been increasingly limited. These restricting constraints have produced bottlenecks in the supply of reagents, consumables, and diagnostic equipment, underscoring the significance of adding additional testing methodologies. Hence, there is a significant need to assess alternative methodologies to ensure that nucleic-acid testing can continue in the face of potential shortages (23-25). In this regard, a molecular approach is the reverse transcription loop-mediated isothermal amplification (RT-LAMP), an amplification method which combines reverse transcription with strand-displacement amplification of DNA in a single reaction at a single temperature (26, 27) thus, eliminating the need of a thermal cycler. RT-LAMP has been applied for the diagnosis of different pathogens in humans, including Zika (28), hepatitis C subtypes (29), human immunodeficiency viruses (30). In the wake of the COVID-19 pandemic, several RT-LAMP protocols have been described (26), including different modifications to improve the sensitivity of the test and the use of colorimetric based detection of the results (31, 32). Applications of RT-LAMP are being evaluated in the clinical setting with favorable results and suggesting that a robust and properly validated RT-LAMP could replace RT-qPCR in specific settings. RT-LAMP might be used as an initial screening tool or in massive population screening, where it can identify highly contagious individuals (26). In addition, the sequencing of SARS-CoV-2 positive samples by RT-LAMP has been considered for a diagnostic validation to detect and record the outcome of RT-LAMP reactions (33).

Since March 2020, our group has been monitoring SARS-CoV-2 genomic dynamics. Variant B.1.1.519 was detected with an epidemic peak that spread throughout the country. Patients infected with variant B.1.1.519 developed symptoms affecting the upper respiratory tract that were associated with an increase in dyspnea (34, 35). However, novel SARS-CoV-2 variants with enhanced infection potential could emerge over time. Therefore, additional molecular tools will be required for the SARS-CoV-2 genomic surveillance, particularly in developing countries where the infrastructure and resources for identifying the virus by using the gold standard test are limited. In this study, we validated the RT-LAMP assay in a mexican cohort that included positive, negative, and inconclusive results in order to determine the RT-LAMP usefulness as a rapid diagnostic method. In addition, our goal was to assess its ability to detect SARS-CoV-2 positive samples, independently of VOC, VOI, VUM and FMV variants that have emerged since the pandemic’s start.

## Material and Methods

### Clinical samples

This cross-sectional, observational study was approved by the Institutional Ethics Committee of the National Institute of Genomic Medicine (INMEGEN) (CEI / 1479/20 and CEI 2020/21). After signing the informed consent, nasopharyngeal swab samples were collected from 984 patients and collected in a 15 ml conical tube with 3 ml of sterile viral transport medium (VTM). After collection, the nasopharyngeal swab samples were processed for viral RNA extraction. 281 SARS-CoV-2 positive patient samples with variants identified by whole genome sequencing were included for independent validation of the RT-LAMP assay.

### SARS-CoV-2 RNA extraction and detection

Viral RNA was extracted from 300 µl of VTM containing nasopharyngeal swabs using the MagMAX Pathogen/Viral Nucleic Acid Isolation Kit and the KingFisher Flex Automated Extraction System (ThermoFisher Scientific) following the manufacturer’s protocol. SARS-CoV-2 RNA detection was carried out with the One-Step RT-qPCR, 5 µl of RNA following the procedures recommended by the CDC (CDC, USA). Two target regions of the N gene (oligonucleotides 2019-nCoV_N1 and 2019-nCoV_N2), are used to detect cases of COVID-19 and human RNase P (RP) as extraction control. Samples were considered as SARS-CoV-2 positive when primer-probe sets N1 and N2 were detected with a Ct less than 40. All tests were performed with the QuantStudio ™ 7 Flex Real-Time PCR System.

#### Oligonucleotide sets

The oligonucleotide sets were designed targeting unique regions of the Spike protein (S) gene, the SARS-CoV-2 RNA-dependent RNA polymerase (RdRP) gene, and fragments of the open reading frame 1a/b (ORF1a/b). Altogether, five sets of LAMP primers were selected for RT-LAMP analysis, where Set 1 and Set 2 targeted the non-structural protein (nsp) 3 region in the ORF1a/b gene and S protein gene, respectively; and Sets 3, 4, and 5 were designed to amplify different regions of the SARS CoV-2 RdRP gene. The oligonucleotides sequences and the first evaluations of each set were previously described by Mohon *et al*., 2020 (36).

#### RT-LAMP assay

Assay validation was carried out in 96-well plates (Applied Biosystems) with a total reaction volume of 20 µL. Briefly, 5.5 µL of molecular biology grade water (Corning), 10 µL of WarmStart Colorimetric RT-LAMP 2X master mix (New England Biolabs), 2 µL of oligonucleotide pool (10x) (S1, S2, S3, S4 and S5), 1 µL of guanidine (20x) (Invitrogen), 0.5 µL of GelGreen 0.07% (Biotium) and 1 µL of RNA was added to each well. The mixture was incubated at 65 °C for 40 min in a PCR thermal cycler (GeneAmp PCR System 9700, Applied Biosystem). The results were determined by visual examination of color modification and by a fluorescence method. Visual inspection was carried out using a transilluminator and fluorescence was detected using a photo documenter (Universal Hood III Gel Documentation System, Bio-Rad), or images were taken with mobile phone cameras (for color evaluation) and fluorescence was visualized and captured with Image Lab 6.1 software (Bio-Rad). The results were interpreted as positive (yellow and fluorescent color) and negative (pink or salmon color and non-fluorescent).

#### Library preparation and sequencing

The libraries were prepared using the Illumina COVID-seq protocol following the manufacturer’s instructions. Briefly, first-strand synthesis was carried out on RNA samples. The synthesized cDNA was amplified using ARTIC primers V3 for multiplex PCR, generating 98 amplicons across the SARS-CoV-2 genome. The PCR-amplified product was tagmented and adapted using IDT for Illumina Nextera UD Indices Set A, B, C, D (384 indices). Dual-indexed single-end sequencing with a 36 bp read length was carried out on the NextSeq 550 platform.

#### Data processing and variant detection

Illumina raw data processing and sequencing data quality assessment. The raw data were processed using DRAGEN Lineage v3.3.4/5 with standard parameters. Further samples with SARS-CoV-2 and at least 90 targets detected were processed for lineage designation.

#### Statistical analysis

The diagnostic utility of the RT-LAMP test for detection of SARS-CoV-2, including analysis of sensitivity and specificity, positive predictive value, and negative predictive value was determined using RT-qPCR as the “gold standard”. Other statistical analyzes were performed using GraphPad Prism 7.0 software and IBM SPSS Statistics version 24. Two-tailed parametric statistical tests (Student’s t test) were used to determine the significance of the data, considering a statistically significant value of p ≤ 0.05. The Kappa coefficient was used to estimate the agreement between the results of RT-LAMP and RT-qPCR.

## Results

### Study population selection

A total of 984 individuals residing in Mexico City were included in this study. General demographic and clinical data was available for 865 patients (88% of the total individuals considered), which allowed determining the clinical condition based on the CDC (Centers for Disease Control and Prevention) clinical guide (37). In addition, we were able to stratify the disease severity in patients with complete clinical data and positive to SARS-CoV-2 by RT-qPCR.

Our study population consisted of male and female patients ranging all ages. We detected, however, a sharp rise in infection cases in people aged 30 to 59 years old. We noted that the majority of individuals had some risk condition, such as heart disease, obesity, diabetes. These chronic degenerative diseases are the most frequent in Mexican population, based on the risk predicted by the population’s comorbidities and have been related to mortality in COVID-19 (38-40). In addition, most of the positive patients with clinical data had moderate symptoms (Table 1).

**Table 1.**
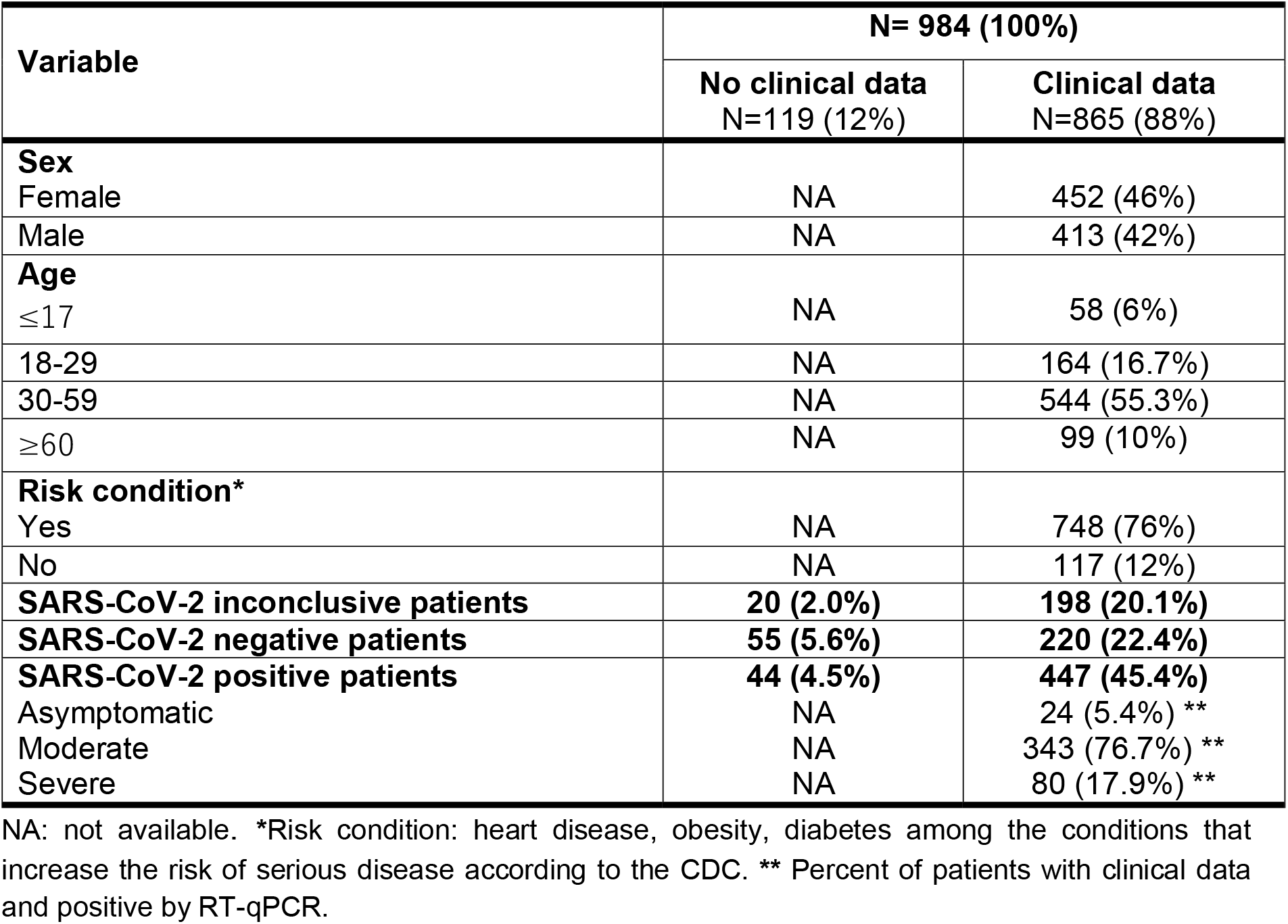
Characteristics of the study population.

| Variable | N= 984 (100%) |  |
| --- | --- | --- |
|  | No clinical data<br>N=119 (12%) | Clinical data<br>N=865 (88%) |
| **Sex** |  |  |
| Female | NA | 452 (46%) |
| Male | NA | 413 (42%) |
| **Age** |  |  |
| ≤17 | NA | 58 (6%) |
| 18-29 | NA | 164 (16.7%) |
| 30-59 | NA | 544 (55.3%) |
| ≥60 | NA | 99 (10%) |
| **Risk condition\*** |  |  |
| Yes | NA | 748 (76%) |
| No | NA | 117 (12%) |
| **SARS-CoV-2 inconclusive patients** | **20 (2.0%)** | **198 (20.1%)** |
| **SARS-CoV-2 negative patients** | **55 (5.6%)** | **220 (22.4%)** |
| **SARS-CoV-2 positive patients** | **44 (4.5%)** | **447 (45.4%)** |
| Asymptomatic | NA | 24 (5.4%) ** |
| Moderate | NA | 343 (76.7%) ** |
| Severe | NA | 80 (17.9%) ** |

NA: not available. \*Risk condition: heart disease, obesity, diabetes among the conditions that increase the risk of serious disease according to the CDC. \*\* Percent of patients with clinical data and positive by RT-qPCR.

#### Evaluation of the sensitivity of oligonucleotides S1-S5

To determine the detection limit and evaluate the sensitivity of the oligonucleotides previously designed by Mohon *et al*., 2020, a standard curve was made for each of the oligonucleotides (S1-S5) by making serial dilutions of 10 factor (1:10, 1:20, 1:30, 1:40, 1:50, 1:60, 1:70, 1:80, 1:90, 1:100 and 1:110), using a positive control (ATCC). We found that the RT-LAMP color detection limit for each of the oligonucleotides is superior (set S1 and S3), since both showed positivity up to the 1:90 dilution (211 RNA copies) (Supplementary Fig. 1). When detected by fluorescence, the sensitivity was up to the 1:50 dilution for all the oligonucleotide sets (Supplementary Fig. 2).

The data results by color and fluorescence, on the other hand, were more consistent with the oligonucleotide sets S1 and S3 (Supplementary Fig. 1 and 2). In addition, we used RT-qPCR to assess the detection sensitivity of RT-LAMP positivity in SARS-CoV-2 positive nasopharyngeal swab samples with CT ranges of 10-15, 16-20, 21-25, 26-30, 31-35, and 36-40. The detection limit of the RT-LAMP test was found to be in the CT range of 26 to 30 by both color and fluorescence (Supplementary Fig. 3 and 4).

The positive and negative samples for SARS-CoV-2 are depicted in Figure 1, together with the standardized parameters of our test. The RT-LAMP test can detect only positive (fluorescent) and negative (non-fluorescent) SARS-CoV-2 clinical samples using the oligonucleotide sets S1 and S3, demonstrating that the RT-LAMP reaction was consistent with negative and positive controls.

**Figure 1.**
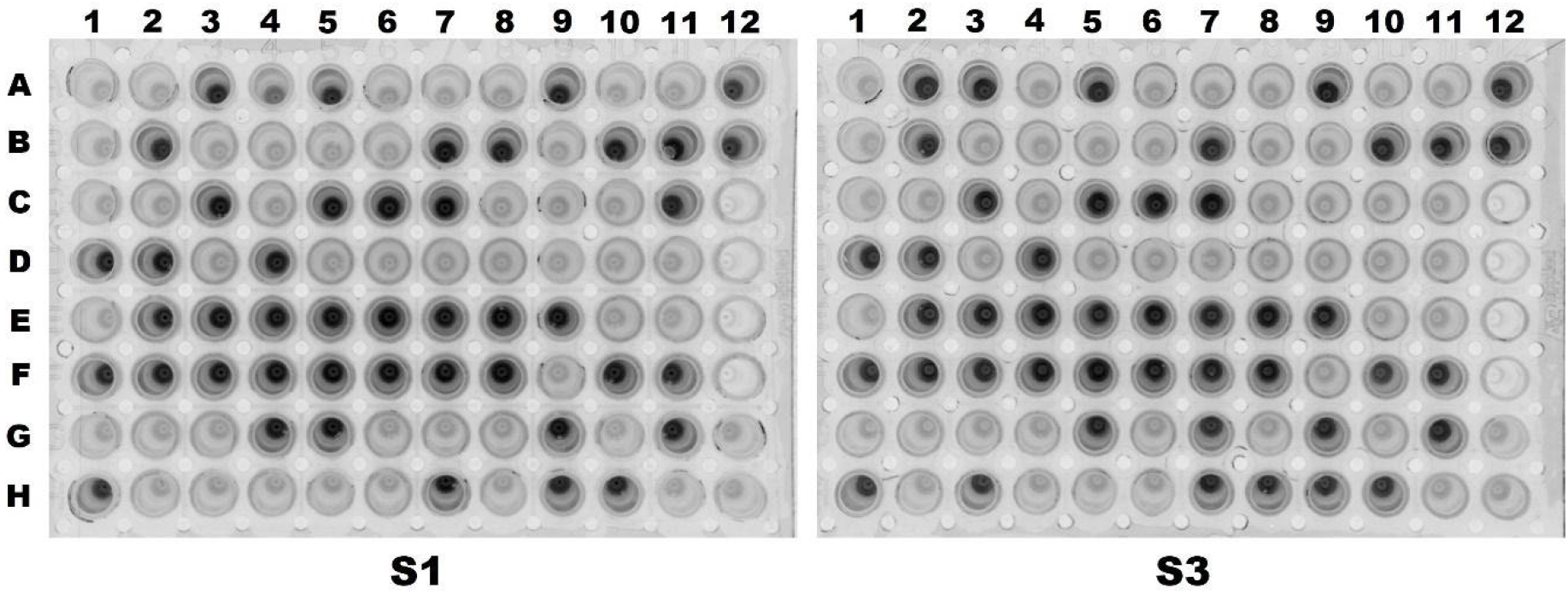
Fluorescence detection of SARS-CoV-2 positive and negative samples using the RT-LAMP assay. The image shows 88 samples processed using the oligonucleotide set **A)** S1 and **B)** S3. The staining of the RT-LAMP products in the 96-well plate were visualized with a photodocumenter. **ATCC positive control:** A12 (stock) and B12 (1:10 dilution); **Negative controls:** G12 (reagents) and H12 (reagents + H_2_O).

#### Validation of the RT-LAMP assay in clinical samples

We performed the RT-LAMP test in 96-well plates with the set of primers S1 and S3 for a massive validation and to determine the efficiency of RT-LAMP in the detection of SARS-CoV-2. We only considered fluorescence-based detection, due to increased sensitivity, to determine positivity and negativity of the assay (Fig. 1A and B). 984 positive, negative, and inconclusive samples of nasopharyngeal swabs with a confirmed diagnosis by RT-qPCR were included in this study, according to the CDC methodology (41) (Fig. 2A). When compared to the set of S3 primers, we observed that the S1 primers detected slightly more positive samples and thus fewer negative samples (Fig. 2B and C).

**Figure 2.**
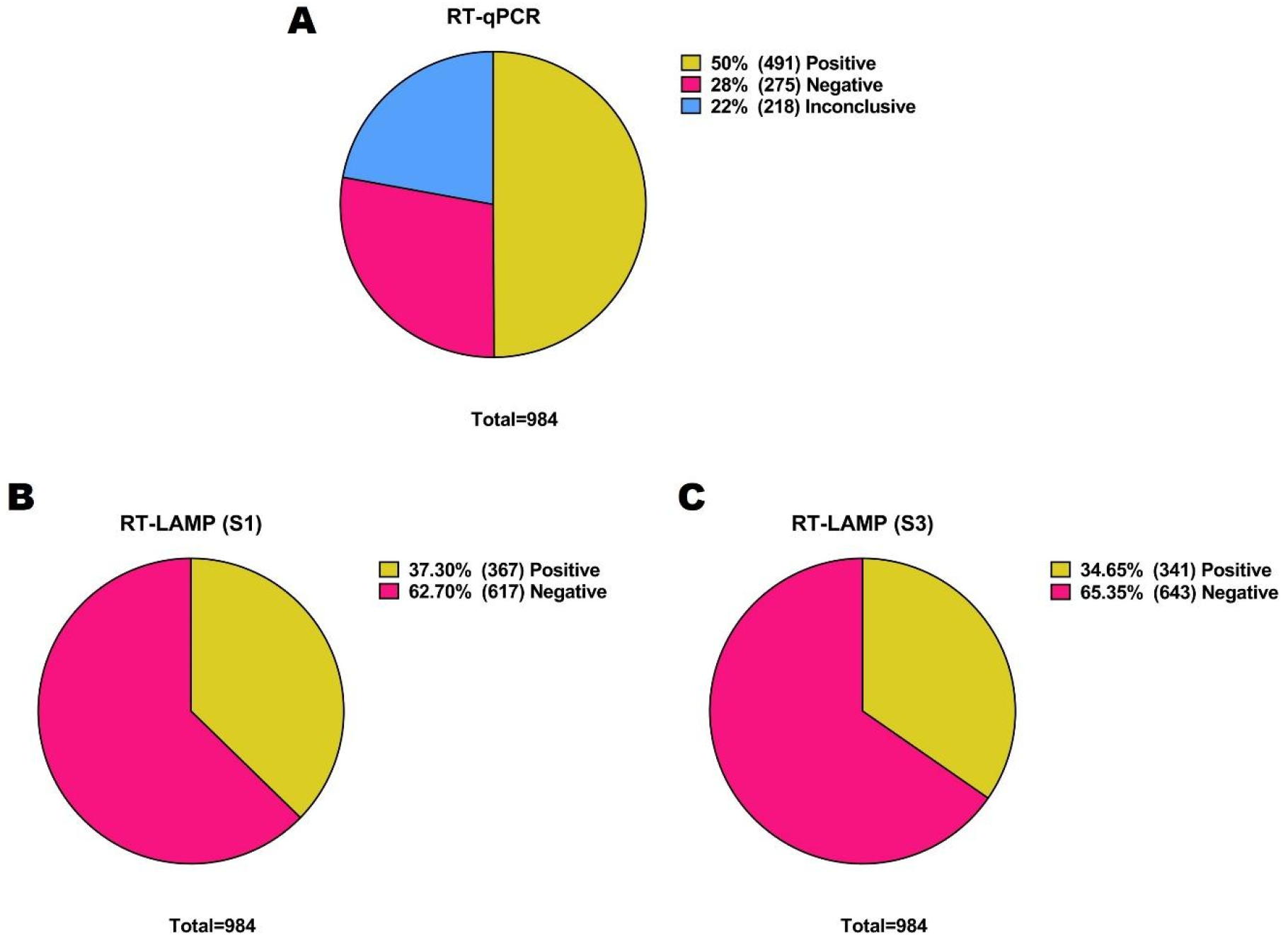
Frequencies and positive/negative percentages of nasopharyngeal swab samples by RT-qPCR and RT-LAMP. **A)** Percentage of positive, negative and inconclusive results determined by RT-qPCR; **B)** and **C)** Percentages of positive and negative results by RT-LAMP of the same samples using the oligonucleotides set to S1 (B) or S3 (C).

#### Comparison of RT-LAMP assay to RT-qPCR CT values

After processing 984 nasopharyngeal swab samples using the RT-LAMP test, we compared the CT values of each of the samples and found that most of the RT-LAMP positive samples with S1 and S3 primers have a CT <35, for both the N1 and N2 genes. However, we observed a group of samples that were amplified at CT <35 that were RT-LAMP negative. We also noticed another group of samples amplified at CT >35 that were RT-LAMP positive using S1 and S3 when plotted with N1 and N2 (Fig. 3A, B, C and D). Thus, we suggest these could represent outliers indicative of the assay’s sensitivity. Furthermore, we expect that using a CT≤35 cutoff point, the RT-LAMP assay can effectively distinguish positive from negative samples.

**Figure 3.**
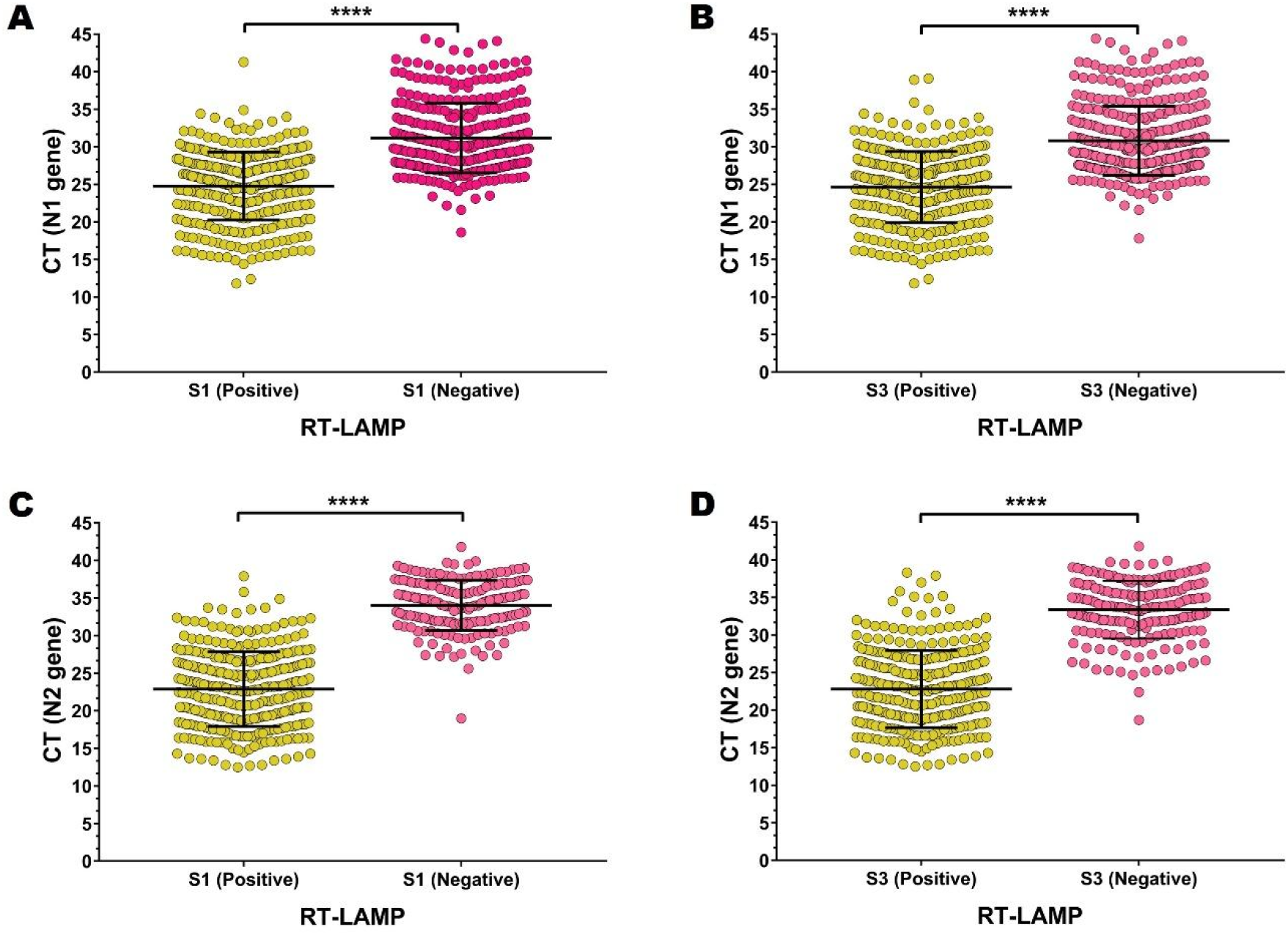
CT values corresponding to all positive or negative results obtained by the RT-LAMP assay. CT values of N1 gene of positive vs negative samples for the set of primers S1 (24.74 ± 0.2337, n=364 and 31.19 ± 0.252, n=339, respectively) **(A)** and S3 (24.63 ± 0.2595, n=333 and 30.8 ± 0.2392, n=370, respectively) **(B)**. CT values of N2 gene of positive vs negative samples for the set of primers S1 (22.89 ± 0.2705, n=337 and 34.03 ± 0.2479, n=181, respectively) **(C)** and S3 (22.81 ± 0.2859, n=323 and 33.37 ± 0.2748, n=195, respectively) **(D)**. **** p<0.0001 (Two-tailed Student’s t-test). P-values ≤0.05 were considered significant.

### Validity of RT-LAMP assay as a diagnostic test for SARS-CoV-2

Using contingency tables, we examined the usefulness of the RT-LAMP test in comparison to the gold standard test (RT-qPCR). We showed that in RT-LAMP, both sets of primers, S1 and S3, correctly identify the majority of inconclusive samples as negative samples by RT-qPCR: 190 (19.31%) and 208 (21.14%), respectively. Furthermore, with both sets of primers (S1: 336/491 and S3: 323/491), RT-LAMP positive samples identified the majority of RT-qPCR positive samples. The RT-LAMP negative samples with both sets of primers, on the other hand, correctly identified virtually all RT-qPCR negative samples (S1: 272/275 and S3: 267/275). (Table 2 and 3). Based on these findings, we suggest that RT-LAMP is more effective at detecting real negatives versus true positives.

**Table 2.**
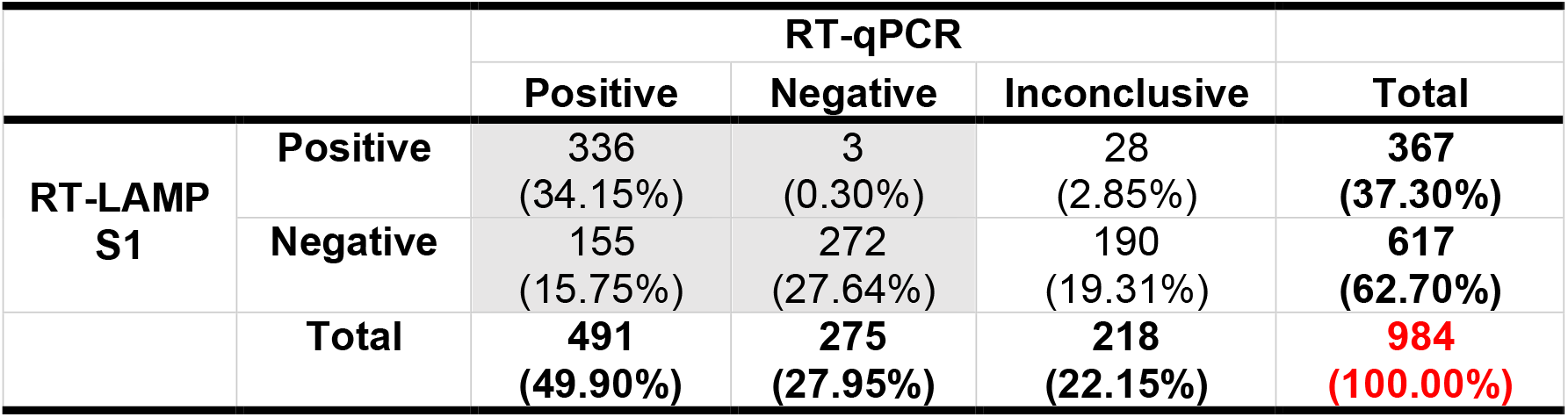
Contingency table of the results obtained with the set of primers S1.

**Table 3.** Contingency table of the results obtained with the set of primers S3.

|  |  | RT-qPCR |  |  | Total |
| --- | --- | --- | --- | --- | --- |
|  |  | Positive | Negative | Inconclusive |  |
| **RT-LAMP**<br>**S3** | **Positive** | 323<br>(32.83%) | 8<br>(0.81%) | 10<br>(1.02%) | **341**<br>**(34.65%)** |
|  | **Negative** | 168<br>(17.07%) | 267<br>(27.13%) | 208<br>(21.14%) | **643**<br>**(65.35%)** |
|  | **Total** | **491**<br>**(49.90%)** | **275**<br>**(27.95%)** | **218**<br>**(22.15%)** | **984**<br>**(100.00%)** |

Similarly, an analysis of sensitivity and specificity, positive predictive value, and negative predictive value was performed using the values true positives (336 for S1 and 323 for S3), true negatives (272 for S1, 267 for S3), false positives (3 for S1, 8 for S3), and false negatives (3 for S1, 8 for S3) to clearly identify the proportion of positive and negative individuals with the RT-LAMP test using the data obtained with the RT-qPCR (155 for S1, 168 for S3).

We observed that the S1 primer set showed marginally higher RT-LAMP sensitivity than the S3 primer set (68.4% and 65.8%, respectively). This could suggest that RT-LAMP has a lower probability of detecting false negatives. Although the sensitivity is moderate, the specificity for both S1 and S3 primers is over 100%, indicating excellent specificity. As a result, the assay is able to detect individuals who are not infected with SARS-CoV-2. The positive predictive value for both S1 and S3 is about 100%, suggesting that the positive results of the RT-LAMP assay correspond to the proportion of subjects who are truly infected with SARS-CoV-2. However, RT-LAMP has a low probability of not detecting those infected with SARS-CoV-2, because of its negative predictive value (Table 4).

**Table 4.** Evaluation of rapid RT-LAMP method for the detection of SARS-CoV-2.

| **Statistical variable** | **S1** | **S3** |
| --- | --- | --- |
| **Sensitivity** | 68.4% | 65.8% |
| **Specificity** | 98.9% | 97.1% |
| **Positive predictive value** | 99.1% | 97.6% |
| **Negative predictive value** | 63.7% | 61.4% |

In addition, we calculated a ROC (Relative Operating Characteristic) curve to represent sensitivity versus specificity and for the interpretation of the ratio or proportion of true positives (TPR = True Positive Rate) versus the ratio or proportion of false positives (FPR = False Positive Rate), using only positive and negative RT-qPCR samples (excluding indeterminate samples). Because the calculated points are above the diagonal with an Area Under the Curve (AUC) of 0.814 and 0.795, respectively, we determined satisfactory categorization or diagnostic results for both sets of oligonucleotides S1 and S3 (Fig. 4). This data suggests that the diagnosis established for a COVID-19 patient has an 81% (S1) and 79% (S3) likelihood of being correct. With these findings, we can suggest that RT-LAMP has a fairly adequate diagnostic value for detecting SARS-CoV-2 when both sets of oligonucleotides are employed.

**Figure 4.**
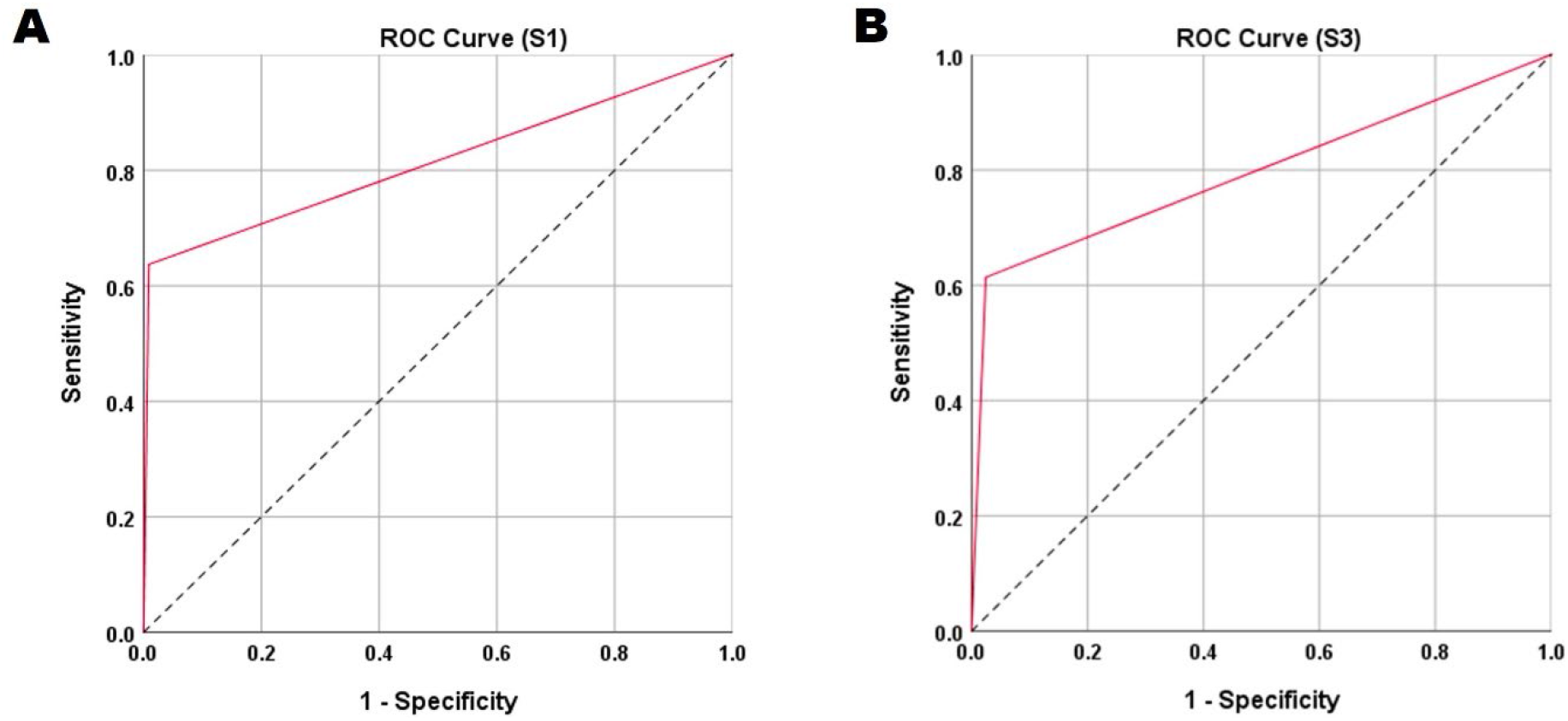
ROC curves for the set of oligonucleotides S1 and S3 used in the RT-LAMP assay. **A)** ROC curve of S1 result: AUC = 0.814, p = 0.000, 95% CI = 0.783-0.845. **B)** ROC curve of S3 result: AUC = 0.795, p = 0.000, 95% CI = 0.763-0.827.

We used the Kappa index concordance analysis on 766 nasopharyngeal swab samples, in order to measure the degree of agreement between the RT-LAMP test and the RT-qPCR. Inconclusive results derived from the RT-qPCR were excluded for this analysis.

We observed that the degree of concordance was statistically significant (p value 0.000 for both oligonucleotides set) and fell between the good (0.6-0.8) and moderate (0.4-0.6) ranges for the S1 (0.600) and S3 (0.557) primer sets, respectively (Table 5). The results of the RT-LAMP replicate the results of the RT-qPCR to some extent, despite the modest agreement.

**Table 5.** Cohen’s kappa for concordance between the diagnosis of SARS-CoV-2 by RT-LAMP and RT-qPCR with the S1 and S3 primers set.

| Primers set | Test | Value | asymmetric standard error | Approx. T | Significance |
| --- | --- | --- | --- | --- | --- |
| **S1** | **Cohen's kappa coefficient** | 0.600 | 0.026 | 18.001 | 0.000 |
|  | **Number of valid cases** | 766 |  |  |  |
| **S3** | **Cohen's kappa coefficient** | 0.557 | 0.027 | 16.852 | 0.000 |
|  | **Number of valid cases** | 766 |  |  |  |

**Table 5.** Variants of SARS-CoV-2 prevalent in Mexico identified by RT-LAMP.

| WHO name | Classification (WHO) | Pango lineage | Total samples | CT | RT-LAMP Result (S1/S3) |
| --- | --- | --- | --- | --- | --- |
| **Alpha** | VOC | B.1.1.7 | 32 | 20.01<sup>†</sup> | Pos/Pos |
| **Beta** | VOC | B.1.351 | 1 | <26 | Pos/Pos |
| **Gamma** | VOC | P.1 | 16 | 20.84<sup>†</sup> | Pos/Pos |
|  |  | P.1.1 | 15 | 20.59<sup>†</sup> | Pos/Pos |
| **Delta** | VOC | B.1.617.2 | 4 | 20.76<sup>†</sup> | Pos/Pos |
|  |  | AY.3 | 8 | 19.79<sup>†</sup> | Pos/Pos* |
|  |  | AY.4 | 17 | 21.11<sup>†</sup> | Pos/Pos |
|  |  | AY.12 | 1 | 16.47 | Pos/Pos |
|  |  | AY.20 | 1 | <26 | Pos/Pos |
|  |  | AY.39 | 1 | <26 | Pos/Pos |
|  |  | AY.100 | 3 | <26<sup>†</sup> | Pos/Pos |
|  |  | AY.126 | 1 | <26 | Pos/Pos |
| **Epsilon** | FMV | B.1.429 | 1 | 25.44 | Pos/Pos |
| **Iota** | FMV | B.1.526 | 8 | 21.63<sup>†</sup> | Pos/Pos |
| **Kappa** | FMV | B.1.617.1 | 1 | 19.09 | Pos/Pos |
| **Lambda** | VOI | C.37 | 24 | 19.67<sup>†</sup> | Pos/Pos |
|  |  | C.37.1 | 1 | <26 | Pos/Pos |
| **Mu** | VOI | B.1.621 | 28 | 18.67<sup>†</sup> | Pos/Pos |
| - | VUM | B.1.1.318 | 2 | 24<sup>†</sup> | Pos/Pos |
| - | FMV | B.1.1.519 | 27 | 19.64<sup>†</sup> | Pos/Pos* |
| - | FMV | B.1.619 | 1 | 20.85 | Pos/Pos |
| - | FMV | B.1.620 | 1 | 18.44 | Pos/Pos |
| **Omicron** | VOC | B.1.1.529 | 1 | <26 | Pos/Pos |
|  |  | BA.1 | 87 | <26<sup>†</sup> | Pos/Pos |

\*Only a negative sample for S1 and S3. <sup>†</sup>CT mean

### Distribution and correlation of the expression of the genes N1 and N2 with the sets of primers S1 and S3

Using the set of primers S1 and S3 and the CT value, we created a distribution plot of positive RT-LAMP test samples. When plotting the N2 gene samples (Fig. 5B), we observed that they had a mean ± SD slightly more dispersed distribution than when plotting with the N1 gene (Fig. 5A). In order to validate if any of the oligonucleotides used in RT-qPCR (N1 and N2) would alter the sensitivity of RT-LAMP, we employed the coefficient of Pearson’s correlation to measure the degree of association between the primers of both SARS-CoV-2 detection procedures.

**Figure 5.**
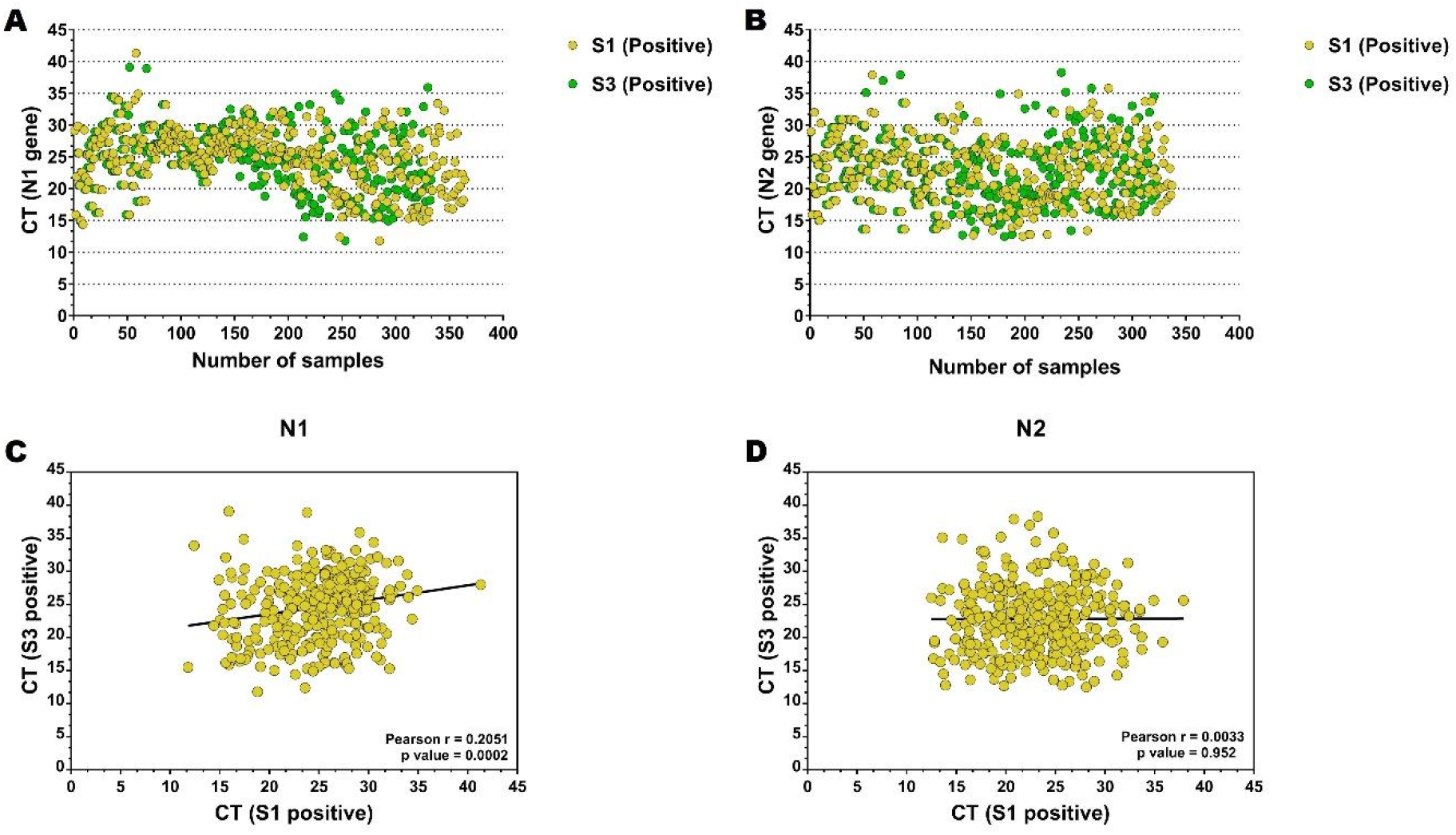
Distribution and correlation of positive samples by RT-LAMP. Comparison of the distribution of positive results obtained by the RT-LAMP test separated by the CT values of **A)** N1 (S1 24.78± 4.53; S3 24.63±4.73) and **B)** N2 (S1 22.89±4.96; S3 22.81±5.13. Pearson’s correlation of the expression of **C)** N1 and **D)** N2 between the positive samples with the set of primers S1 and S3. Pearson’s correlation coefficient (r) and p-values are shown for each analysis. P = values ≤0.05 were considered significant.

SARS-CoV-2 detection using the set of primers S1 and S3 shows a shallow but statistically significant positive correlation with the detection of the N1 gene (Fig. 5C), however there is no significant association when S1 and S3 are compared to the detection of the N2 gene (Fig. 5D).

This suggests that more adequate data could be obtained from RT-LAMP only by considering the N1 gene results. This can be observed when plotting the N1 and N2 derived data (Supplementary Fig. 5), where oligonucleotide N1 detects more positive samples and fewer indeterminate samples than oligonucleotide N2.

### An independent validation with RT-LAMP detects SARS-CoV-2 positive samples with VOC, VOI, VUM and FVM variants

In order to further validate the RT-LAMP assay, we analyzed SARS-CoV-2 genome sequencing results in an independent cohort of 281 positive patients to identify variants with different lineages of clinical interest (VOI), Variants of Concern (VOC), Variants Under Monitoring (VUM), and Formerly Monitored Variants (FVM) (classification according to WHO, 17 January 2022) (42). We found that the RT-LAMP assay adequately identifies all samples with Alpha, Beta, Gamma, Delta, and Omicron VOC variants (Fig. 6A and B; Table 5), as well as VOI, VUM and FVM (Fig. 6C and D; Table 5) (sample data is shown in Supplementary Table 1). This independent validation cohort is composed of SARS-CoV-2 positive samples, validated by RT-qPCR and sequenced with CT <29. Only two samples failed to be detected by RT-LAMP.

**Figure 6.**
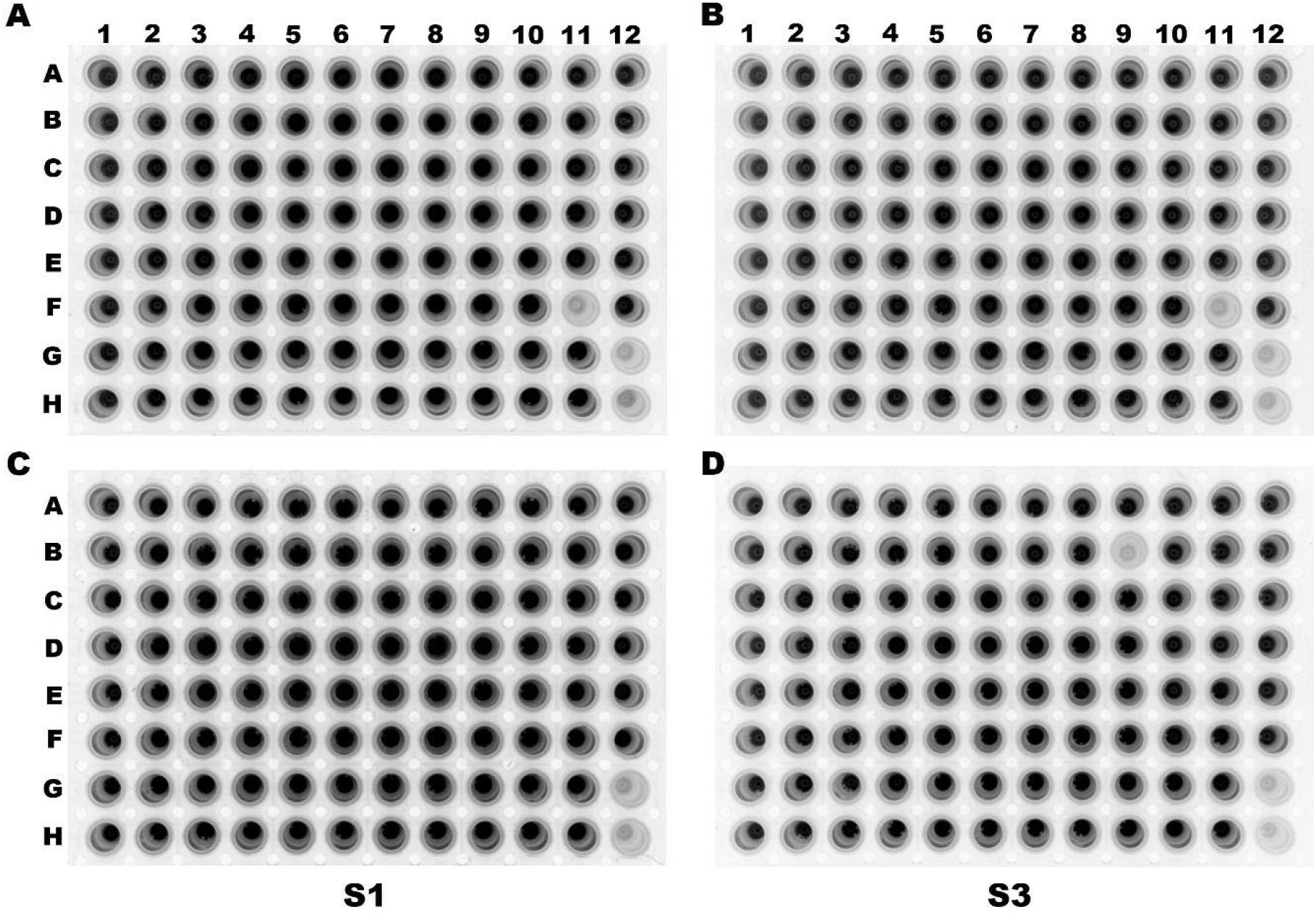
Validation of the RT-LAMP assay with SARS-CoV-2 variants. VOC variants **A)** S1, **B)** S3; VOI, VUM and FMV variants **C)** S1 y **D)** S3. The staining of the RT-LAMP products in the 96-well plate were visualized with a photodocumenter. Negative controls: G12 (reagents) and H12 (reagents + H_2_O).

Finally, we determined the sensitivity of the RT-LAMP assay with positive samples with different variants that have appeared throughout the pandemic in Mexico City. We found the sensitivity to be excellent (99.3% for both S1 and S3) (Table 6 and 7). Taken together, our data suggests that RT-LAMP positive samples are potential candidates for viral genome sequencing and subsequent genetic variant determination.

**Table 6.** Contingency table of the results obtained with the set of primers S1 of sequencing samples.

|  |  | RT-qPCR |  |
| --- | --- | --- | --- |
|  |  | Positive | Total |
| **RT-LAMP S1** | **Positive** | 279<br>99.3% | 279<br>99.3% |
|  | **Negative** | 2<br>0.7% | 2<br>99.3% |
|  | **Total** | **281**<br>**(100.00%)** | **281**<br>**(100.00%)** |

**Table 7.** Contingency table of the results obtained with the set of primers S3 sequencing samples.

|  |  | RT-qPCR |  |
| --- | --- | --- | --- |
|  |  | Positive | Total |
| **RT-LAMP**<br>**S3** | **Positive** | 279<br>99.3% | 279<br>99.3% |
|  | **Negative** | 2<br>0.7% | 2<br>99.3% |
|  | **Total** | **281**<br>**(100.00%)** | **281**<br>**(100.00%)** |

## Discussion

In this study, we validated the performance and diagnostic utility of RT-LAMP for the detection of SARS-CoV-2 in the largest prospective cohort of positive samples to test such an assay. In addition, we include negative and inconclusive samples confirmed by RT-qPCR in order to establish a quality control for each batch of reagents and to determine whether the conditions used in the processing of the samples are capable of reproducing the results obtained by the gold standard (43), as the reaction conditions of the reagent, the reaction system, and the amount of nucleic acid addition can affect the sensitivity of detection and analysis (44). First, we tested five set of oligonucleotides (S1, S2, S3, S4, and S5), designed by Mohon and collaborators (2020) (36) using a ATCC positive control and SARS-CoV-2 positive samples with different CTs and we obtained the most satisfactory results with the sets S1 (designed in the ORF1a/b (nsp3) gene) and S3 (designed in the RdRP gene), since they showed more sensitive and consistent results. Additionally, the set S1 seems to be slightly more sensitive than the set S3 for the detection of SARS-CoV-2 infection. However, we consider that both sets are useful for the RT-LAMP assay, since oligonucleotides targeting regions of the *RdRP* and *ORF1a/b* genes are employed in the RT-qPCR methods used to identify SARS-CoV-2, which have shown to be highly sensitive (up to 100%) (45-51). Furthermore, because the ORF1a/b gene is a conserved genomic region of the virus, it has been the target of oligonucleotide design (48, 49), and thus, we presume that the set S1 will perform slightly better than the set S3 (52, 53). The colorimetric analysis, on the other hand, did not yield positive results, probably due to the sample extraction conditions. Similarly, several factors, including the viral RNA elution buffer, which is compatible with the RT-LAMP reaction but could dramatically impact colorimetric readings, have been noted as potentially interfering with the pH change (54). High concentrations of other RNA types, such as rRNA, also might interfere with the results. Therefore, we used fluorescence to validate RT-LAMP.

We then used the RT-LAMP test to evaluate the detection limit of SARS-CoV-2 based on CT value and we observed that, similar to previously reported, the cut-off point in our study corresponded to CT values of 35. In one report, the RT-LAMP detection limit for SARS-CoV-2 detection corresponded to CT values of 34.2 employing SARS-CoV-2 RNA isolated from VERO cells, whereas 14 positive samples by the RT-LAMP test of 154 clinical samples had CT values ranging between 21.11 to 32.76 (55). A highest value of CT 36.64 was reported for the identification of SARS-CoV-2 by RT-LAMP in another study that included 16 samples (8 positive and 8 negative) (54).These data indicates that our findings are consistent with previously published research evaluating SARS-CoV-2 in nasopharyngeal swab samples. According to our assessment of the RT-LAMP assay’s diagnostic usefulness, we observed an increased proportion of real positive results than false positive results, although a better proportion of true negatives. Interestingly, the RT-LAMP assay classified the inconclusive RT-qPCR samples as negative, possibly due to the extremely low or non-existent viral load in these samples, since most of them only amplified one of the primers (N1 or N2) and had a CT of 36 for the RT-LAMP result. This was confirmed in the sensitivity of validation of the RT-LAMP assay. However, we did not expect a higher sensitivity than RT-qPCR, which is the gold standard for SARS-CoV-2 detection due to its excellent sensitivity and specificity (45, 56-58). Nevertheless, it has been suggested that high sensitivity values could act as a double-edged sword, due to cross-contamination in RT-LAMP reactions, which can lead to false-positive results (54, 59, 60).

Furthermore, the sensitivity of RT-LAMP is comparable with commercial rapid tests, based on the qualitative detection of specific SARS-CoV-2 antigens such as Roche. Due to the fact that the sensitivity of this test declines with later CTs, it was classified as moderate (95%; CT 25 to 30), low (44.8%; CT 30 to 35) and very low sensitivity (22.2%; CT > 35) (61). Other studies describe this test with sensitivity of 61.5% (62), 66.3% (63), up to 71.43% (64). Rapid tests, such as the Abott CLMSRDL, and DIALAB, show a sensitivity between 45.2% and 88.9%, respectively. However, these tests display a significant diagnostic variation, which increases the possibility of not detecting infected people (65). The type of sample is another aspect that influences the assay’s sensitivity. It has been reported that the quality and amount of viral RNA in COVID-19 molecular testing samples is significantly dependent on the type of samples taken (66, 67). Thus, the detection rate of SARS-CoV-2 by RT-qPCR in patients is higher in bronchoalveolar lavage fluid (93%) than in sputum and nasopharyngeal swab samples (72% and 63%, respectively) and in uncommon samples such as pharyngeal smears and feces (32% and 29%) (68). Even using saliva and direct swab to the RT-LAMP assay, it shows variability in the sensitivity of the method, where sensitivity for direct RT-LAMP on saliva was in general higher than that determined for swabs (CT <33=87.61%, CT <45=84.62%) (69). Nevertheless, it has been suggested that determining the absolute sensitivity of the current tests used to detect SARS-CoV-2 (RNA genomes per milliliter) is challenging, as even the gold standard is not definite for the detection of a pathogen that has only been known for a small period and varies constantly. This is why it’s important to underline that “bad but cheap” tests can be diagnostically useful, assuming that the tests’ limitations are carefully evaluated (70, 71).

In our study, however, the specificity of RT-LAMP was 98.9% for the S1 oligonucleotide set and 97.1 % for the S3 oligonucleotide set, indicating that the test is an excellent method for detecting SARS-CoV-2 despite the presence of other interfering molecules isolated during RNA extraction from nasopharyngeal swab samples. Furthermore, based on the positive and negative predictive value of our data, the RT-LAMP assay could be used for massive COVID-19 screening.

As a consequence, the strong positive predictive value (99.1 % for set S1 and 97.6 % for set S3, respectively) indicates that patients who have a positive RT-LAMP test actually have the condition; whereas the negative predictive value for both oligonucleotide sets was modest, this suggests that even if the test was negative, there is still a risk of infection. As a result, we suggest that using a ROC curve analysis to directly compare the cost/benefit of the RT-LAMP assay and other diagnostic procedures is appropriate for making diagnostic decisions.

Although, other study describes greater sensitivity results for the RT-LAMP assay (20 RNA copies per reaction), comparable to RT-qPCR test (72). In this regard, we suggest that the small number of samples and the design of primers based on only 130 fully aligned SARS-CoV-2 genomes are important limiting factors in determining the sensitivity and specificity of any assay, particularly when it is used to evaluate a test as a diagnostic method for the analysis of large numbers of tests. The authors discuss a critical point, claiming that mutations in the region of the target gene’s primer sequence affect the precision of this RT-LAMP assay. Therefore, they suggest that it is necessary to monitor these mutant sites by sequencing the whole virus genome (72). Considering these previous data, we sequenced 281 SARS-CoV-2 whole genome samples for validation of the RT-LAMP assay. This validation analysis included different variants circulating in Mexico City, named VOC, VOI, VUM and FVM by the WHO (42). Validation analysis helped us to determine if the results of the RT-LAMP were affected by variants, which we identified by whole genome sequencing. We found that RT-LAMP detected 279 positive samples out of 281 sequenced samples with different variants with an excellent sensitivity of 99.3% for both sets of oligonucleotides. Therefore, we suggest that RT-LAMP positive samples are potentially candidates for virus sequencing and subsequent identification of SARS-CoV-2 genomic variants, since the validated protocol for the sequencing of SARS-CoV-2 includes positive samples with CT <35 (73).

Preliminary results showed that a few positive samples by RT-LAMP test (CT ≤34.9) were efficiently sequenced with Oxford Nanopore technology (LamPORE). This aims to genotype the virus and, in the future, provide an alternative approach for detecting SARS-CoV-2 and performing genomic surveillance both in symptomatic and asymptomatic individuals, even employing a variety of samples (74-77). Unfortunately, few studies report RT-LAMP validation in samples based on variants propagated in Vero cells (78), or the RT-LAMP assay is focused on validation with a VOC variant (79). We propose that validating and using RT-LAMP to identify infected patients could make this assay a very important tool for epidemiological and genomic surveillance of SARS-CoV-2 in laboratories where the necessary infrastructure is not available, even in saliva samples that have turned out to be a useful type of sample for the diagnosis of SARS-CoV (80).

Finally, the sensitivity of RT-LAMP depends on an appropriate oligonucleotide design, the number of viral RNA copies in the sample and the sample type used for the assay. Furthermore, the validation studies of molecular tests, mainly RT-LAMP, for SARS-CoV-2 detection, generally display limitations such as the small number of samples; differences in the sample collection, storage, and manipulation before diagnostic testing (preanalytical bias); and lack validation by independent laboratories. Additionally, many studies use dispersed clinical parameters, which could hinder the development of diagnostic tests during coronavirus outbreaks (81). Thus, we validated the RT-LAMP molecular test in the largest cohort of positive nasopharyngeal swab samples. We acknowledge that this assay is flexible, low-cost and accessible, which can in turn result advantageous for the detection and monitoring of SARS-CoV-2. Furthermore, despite its good sensitivity and specificity, no diagnostic test has a sensitivity and specificity of 100%, so it should be taken into consideration when the diagnostic results are interpreted in clinical practice (71).

## Conclusion

In conclusion, the RT-LAMP assay could be suitable for screening SARS-CoV-2 infection in suspected patients, especially in clinical laboratories with limited equipment and resources. Furthermore, this assay is another effective molecular test for the control of the SARS-CoV-2 sickness, particularly for the confinement of positive patients using this technology and preventing the spread of the disease, especially in remote locations where lab capacity is limited. Furthermore, each country’s strategy for using alternative rapid tests should consider that no trained personnel is required, and it is useful to initially stratify patients based on positive and negative results. Finally, to confirm the diagnosis resulting from a low viral load and not detected by RT-LAMP, a more sensitive and specific test, such as RT-qPCR, still will be required.

## Data Availability

All data produced in the present work are contained in the manuscript.
All data from sequenced samples included in this study is available at GISAID Initiative web page.

## Acknowledgements

The authors acknowledge the work of the personnel who worked in the SARS-CoV-2 molecular diagnostic laboratory at INMEGEN for the processing of the samples used for this study.

## Data availability

All data from sequenced samples included in this study is available at GISAID Initiative web page (https://www.gisaid.org/, accessed on 18 January 2022).

## Ethics declarations

The study was conducted according to the guidelines of the Declaration of Helsinki and approved by the Institutional Ethics and Research committees of Instituto Nacional de Medicina Genomica (protocol code CEI/1479/20 and CEI 2020/21). Informed consent was obtained from all subjects involved in the study.

## Supplementary Data

**Supplementary Figure 1.**
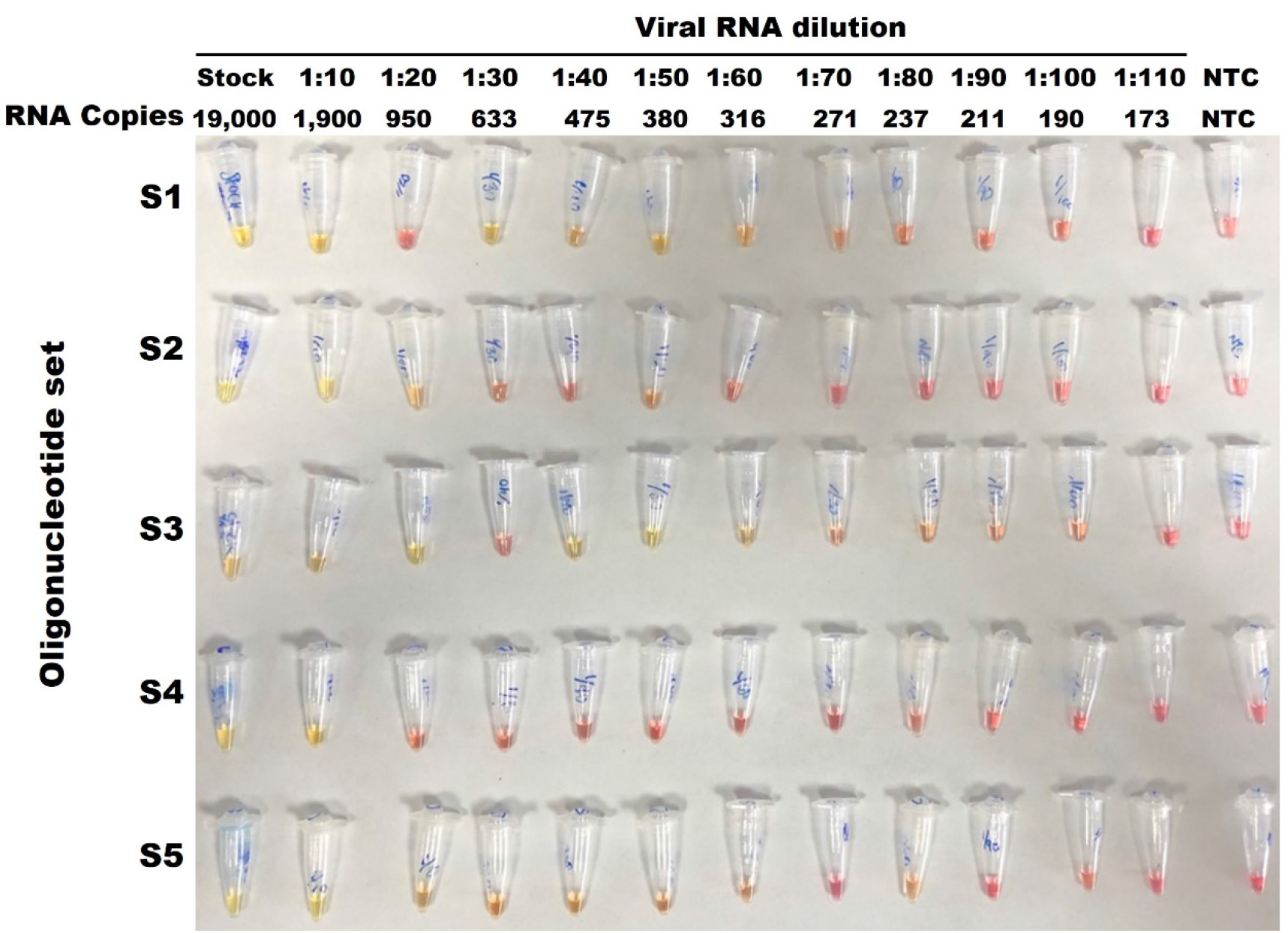
Detection limit of SARS-CoV-2 RNA by RT-LAMP assay detecting by color shift. The image shows the representative results of the standard curve for detection of purified SARS-CoV-2 RNA (ATCC) in a serial dilution (1:10 to 1: 110), by means of an RT-LAMP test, using the oligonucleotide sets S1, S2, S3, S4 and S5. The positive detection of the reaction was determined by the change from pink (negative) to yellow (positive). The number of RNA copies present in each RT-LAMP reaction is shown, corresponding to each dilution. NTC: Non-Temple Control.

**Supplementary Figure 2.**
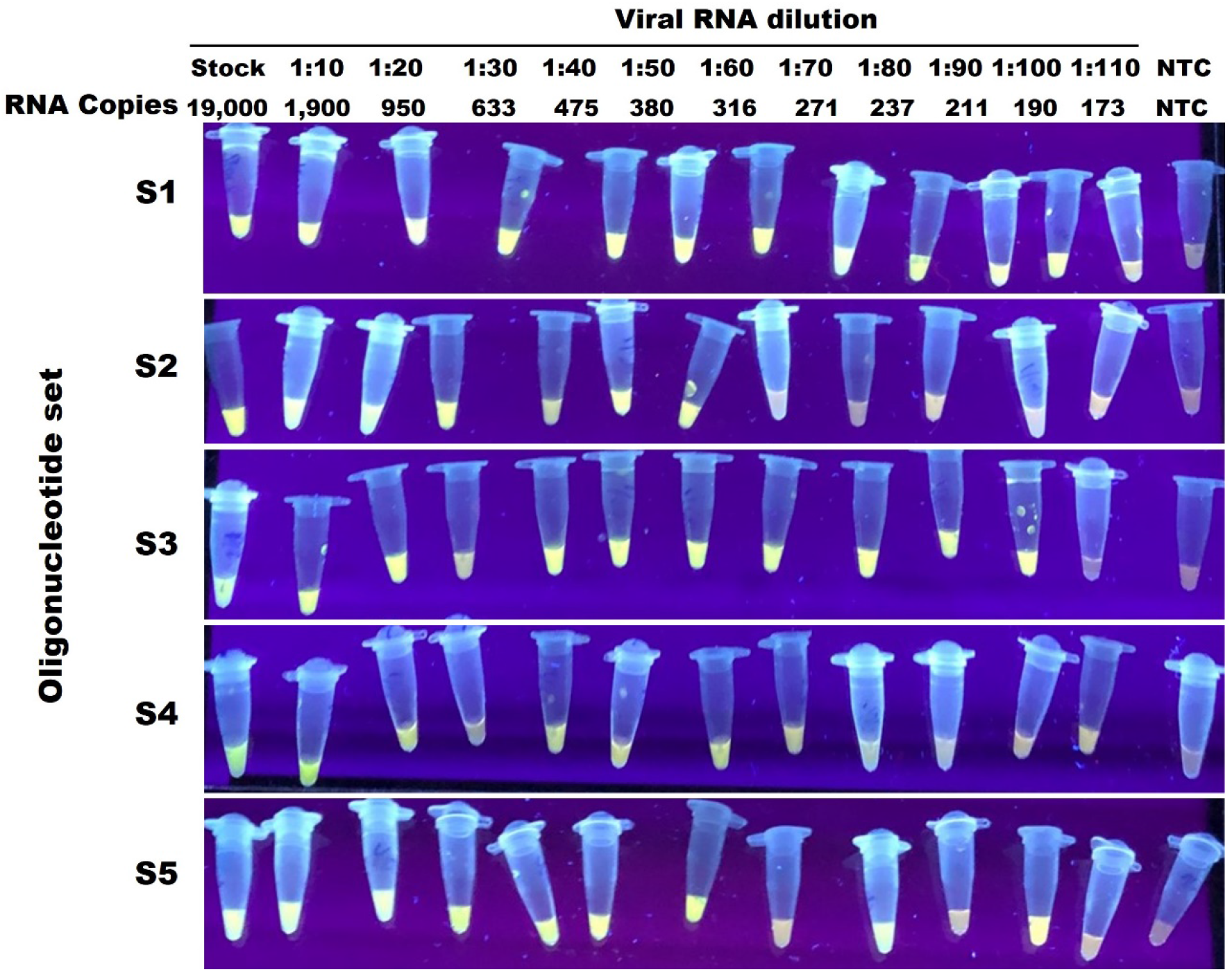
Detection limit of SARS-CoV-2 RNA by RT-LAMP assay detecting by fluorescence. The images show the representative results of the standard curve for detection of purified SARS-CoV-2 RNA (ATCC) in a serial dilution (1:10 to 1: 110), by means of an RT-LAMP test, using the oligonucleotide sets S1, S2, S3, S4 and S5. The positive detection of the reaction was determined by presence of fluorescence (positive). The number of RNA copies present in each RT-LAMP reaction is shown, corresponding to each dilution. NTC: Non-Temple Control.

**Supplementary Figure 3.**
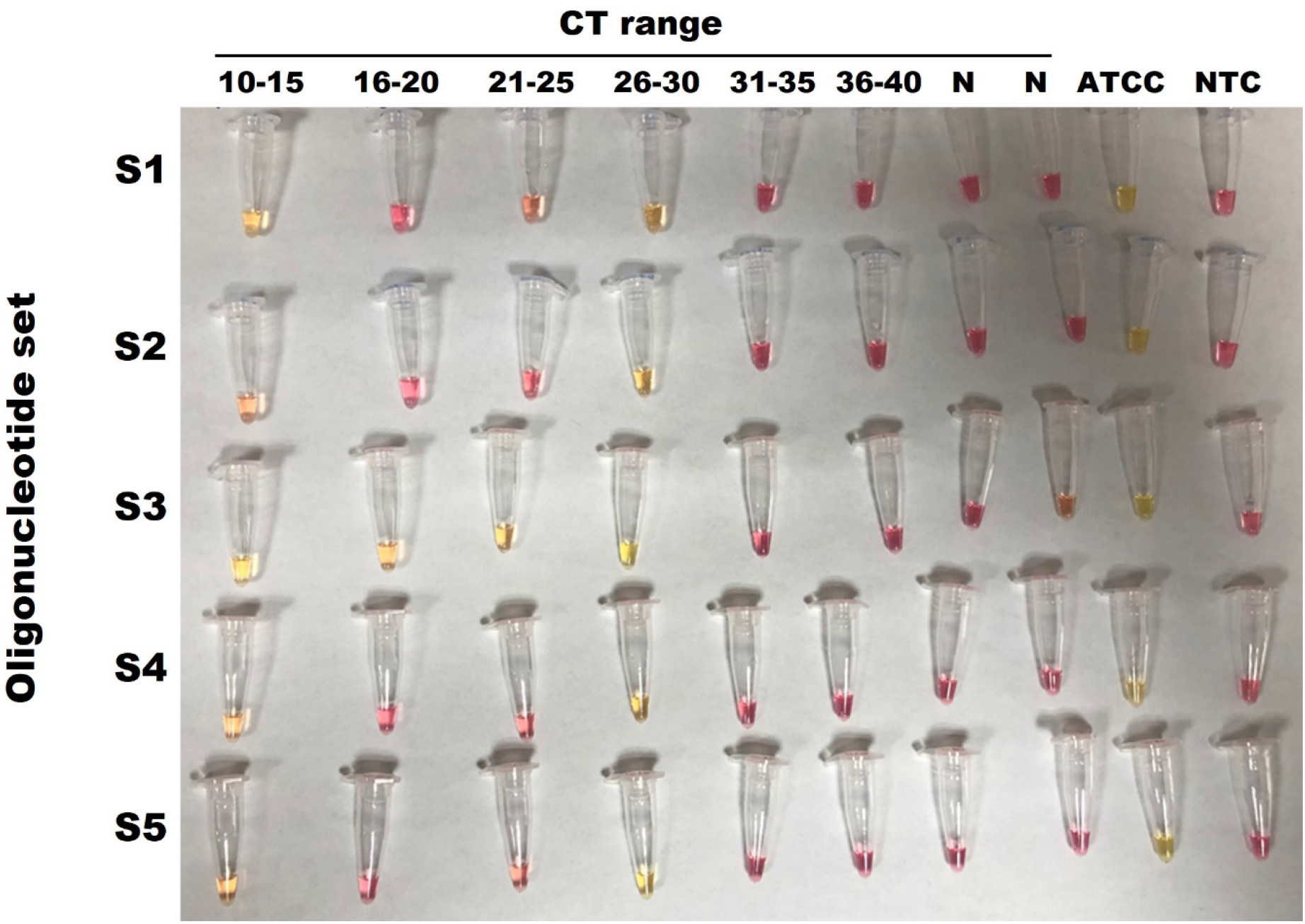
Detection limit of SARS-CoV-2 in clinical samples with different CT values by RT-LAMP assay detecting by color shift. The image shows the representative results of RT-LAMP test to SARS-CoV-2 detection in nasopharyngeal swab with different CT values of N1 and N2 genes (determined by RT-qPCR). The SARS-CoV-2 detection was using the oligonucleotide sets S1, S2, S3, S4 and S5. The positive detection of the reaction was determined by the change from pink (negative) to yellow (positive). N: negative samples by RT-qPCR; ATCC: positive control; NTC: Non-Temple Control.

**Supplementary Figure 4.**
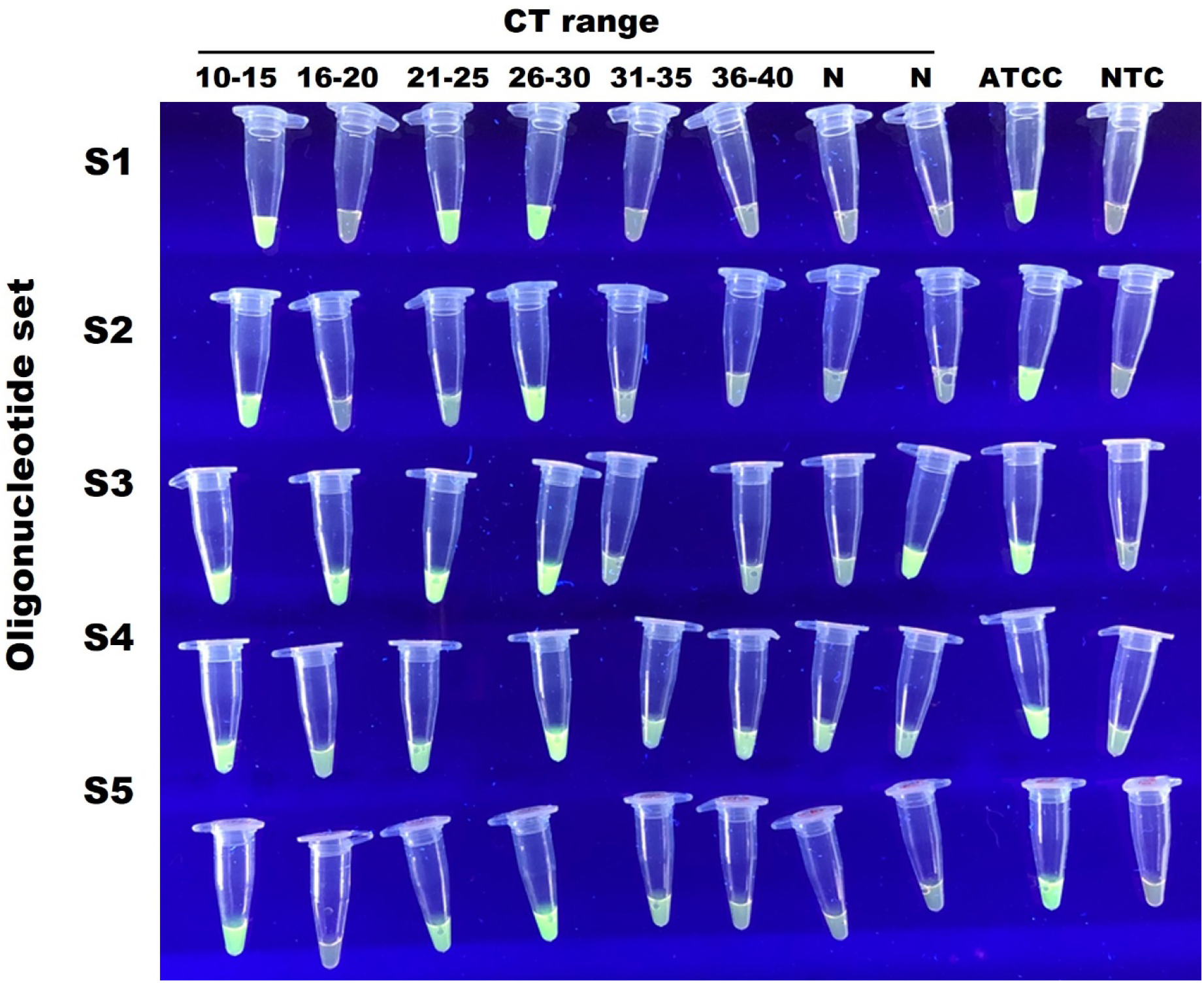
Detection limit of SARS-CoV-2 in clinical samples with different CT values by RT-LAMP assay detecting by fluorescence. The image shows the representative results of RT-LAMP test to SARS-CoV-2 detection in nasopharyngeal swab with different CT values of N1 and N2 genes (determined by RT-qPCR). The SARS-CoV-2 detection was using the oligonucleotide sets S1, S2, S3, S4 and S5. The positive detection of the reaction was determined by presence of fluorescence (positive). N: negative samples by RT-qPCR; ATCC: positive control; NTC: Non-Temple Control.

**Supplementary Figure 5.**
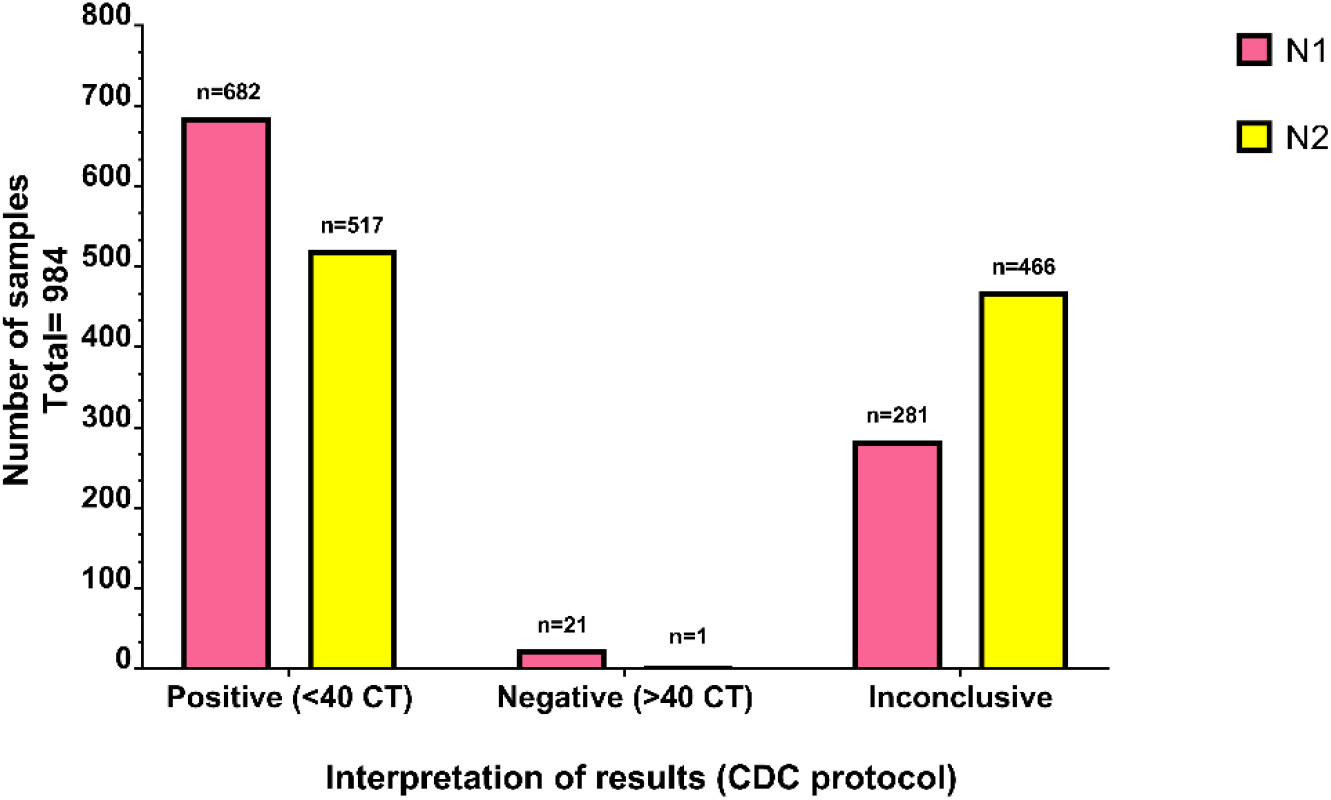
Distribution of positivity of clinical samples determined by amplification of the N1 or N2 genes. Number of positive, negative, and inconclusive samples of nasopharyngeal swab, determined by CT < 40 (positive) or CT > 40 (negative) by RT-qPCR.

